# Impact of COVID-19 on the cascade of care for tuberculosis: A systematic review

**DOI:** 10.1101/2023.07.09.23292326

**Authors:** Tomiwa Fapohunda, Lovemore Mapahla, Reham Amin Khidir Taha, Tawanda Chivese

## Abstract

**Objectives:** To describe the impact of the COVID-19 pandemic on the TB care cascade.

**Methods:** In this systematic review, the Cochrane library, Scopus, CINAHL, Ebscohost, and PubMed databases were comprehensively searched from the onset of the pandemic, till May 5^th^, 2022. Eligible studies were those reporting on changes in the TB cascade of care one year before and one year during the COVID-19 pandemic. Due to the expected differences in the contexts of the included studies, a narrative synthesis was conducted.

**Results:** Twenty-seven studies, from Asia, North America, Africa, South America, and Europe were included. TB screening decreased by between 1% - 50%, and multidrug resistance tuberculosis (MDR-TB) screening decreased by between 15%-17%. Diagnostic delay increased by between 35 - 45 days, contact tracing decreased by up to 36.1%, and case notification decreased by between 3%-63%. TB treatment enrolment decreased by between 16%-35.0%, treatment completion decreased by around 8.0% and treatment success decreased by up to 17.0%.

**Conclusion:** COVID-19 had a detrimental impact on the TB care cascade and these findings suggest a need for policies to protect healthcare systems for TB and other communicable diseases in future health emergencies.

Protocol registration - PROSPERO: CRD42021272456

**Ethics approval:** This systematic review used data from published studies and aggregated data, thus, ethics approval was not required.

## 1. Introduction

Tuberculosis, (TB) remains one of the deadliest infectious diseases in the world, with about 1.8 billion persons infected, 10.6 million ill with TB, and 1.5 million deaths due to TB in the year 2018, most deaths being in the high-burden TB countries [1–3]. The disease has an outsized impact in the countries with the high-burden TB countries, namely; Indonesia, India, China, Pakistan, the Philippines, Nigeria, South Africa and Bangladesh, which are also in the low-to-middle-income (LMIC) bracket [1]. In 2014, the 67^th^ World Health Assembly endorsed the End TB strategy, which aimed for a TB-free society by 2035 [4]. The End TB strategy aimed to reduce TB deaths and incidence by 90% and 80%, respectively, and eliminate the catastrophic costs of the affected households by 2030 [4]. Several countries were taking steps to achieve this but the progress may have been delayed and perhaps, reversed in some cases, when the COVID-19 pandemic began [5]. However, the true impact is still not very clear.

In many countries, the initial policy responses to the COVID-19 included restricted movement and lockdowns [6]. This negatively impacted the healthcare service delivery, especially access to care for people with pre-existing illnesses [7]. Indeed, several countries shifted policies and priorities to combating the COVID-19 pandemic, and, in many cases, at the expense of other health conditions [2]. At certain times, healthcare systems in countries such as Germany, US, Italy, India, and United Kingdom (UK) were overwhelmed with the COVID-19 pandemic management that hospital bed space did not accommodate all the affected individuals, causing them to turn away patients with pre-existing diseases (7). Even though policy responses were different in different countries, in terms of intensity and timing, the interconnected nature of the global healthcare economy meant that many countries were affected by, for example, restrictions in exports of healthcare products in a severely affected country. In low-to-middle-income countries (LMICs), with vulnerable health systems, the pandemic and its associated public health measures may have not only impacted healthcare service delivery but entire health systems [8].

The TB cascade of care was especially vulnerable to disruption as it requires contact between care workers and infected individuals during each of the care stages, from screening to treatment. The TB care cascade is a model of care for the sequential progression of infected individuals from screening, testing and diagnostics, until successful treatment of the disease [9]. The care cascade comprises of screening and testing, diagnosis and confirmation of active TB, TB notification, treatment onset, treatment completion, and post-treatment of individuals with TB [10]. The care cascade concept was adapted from HIV programs and is used to evaluate health care delivery, for programmatic evaluations, and to evaluate the effect of health system interventions [11]. The TB care cascade has also been incorporated in national strategic plans in some high burden TB countries such as India and South Africa [11]. The cascade is designed to ensure positive outcomes such as treatment completion, recovery and the avoidance of disability and death, but interruption of the cascade may have negative outcomes such as TB recurrence, MDR and XDR-TB, incomplete treatment, relapse, re-treatment, and death [10]. In many countries, the COVID-19 pandemic may have disrupted some of the components of the cascade, not only through the public health measures against COVID-19 but also through re-deployment of experienced primary care, laboratory, respiratory and allied health professionals to the care COVID-19 infected individuals. The impact was worsened by the physical and mental demands that were placed on healthcare workers, in the face of a fast-spreading disease that had no cure, which resulted in the death of many key healthcare personnel, especially before the introduction of vaccines [12]. Not only were human resources diverted to the COVID-19 response, but TB infrastructure, due to the similarities of the two respiratory diseases, was also used. For example, centres for TB management in some countries were changed to COVID-19 testing and treatment centres [13]

Several studies [14–17] have documented the effects of the COVID-19 pandemic on some components of the TB cascade of care but estimates of the effect of COVID-19 vary, perhaps because of the different ways that the countries were affected by COVID-19 and how and when they responded. For example, a very small (1%) decrease in numbers screened for TB from 12 months prior to COVID-19 was reported in Vietnam [14], while in India, TB screening went down by 50% [14]. This research investigated the effect of the COVID-19 pandemic on the TB care cascade, through a systematic synthesis of findings from existing research. Specifically, this study compared TB screening, notification, and treatment before and during the COVID-19 pandemic.

## 2. Methods

### 2.1. Study design

The design and methods of this systematic review were based on the Preferred Reporting Items for Systematic Reviews and Meta-Analysis (PRISMA) guidelines [18]. The protocol of the systematic review is registered on the International prospective register of systematic reviews (PROSPERO) with registration number CRD42021272456.

### 2.2. Information sources

The COCHRANE Library, Scopus, CINAHL, Ebscohost, and PubMed databases were searched, without language restriction, and the references of each included study were also searched manually.

### 2.3. Search strategy

The database search was from December 1^st^, 2019, to October 1^st^, 2021, and an updated search was conducted from September 1^st^, 2021, to May 5^th^, 2022. The full search strategy and terms are shown in Supplementary Table 1.

### 2.4. Study selection and eligibility

The study records from the searches were exported to Endnote referencing software for duplicate removal and then exported to the Rayyan systematic review management website (https://www.rayyan.ai/) for initial screening using the title and abstract screening. Two reviewers conducted the screening independently, any conflicts were addressed via consensus, and a third reviewer resolved discrepancies when consensus was not reached. After the initial selection, full text assessment of eligibility was carried by two independent authors, using predefined eligibility criteria.

Studies were included if they were observational studies such as cohort, cross-sectional, case series, interrupted time series and population-based studies that quantitatively described the number or percentage change in any of the outcomes; TB screening, case notification, diagnosis, and treatment at least one year before and one year during the pandemic. Qualitative studies, reviews, case studies, letters to the editor and commentaries were excluded.

### 2.5. Data extraction

Two reviewers extracted data from each study independently, and a third reviewer resolved any disagreements. We extracted data on characteristics of studies such as study title, authors, years of publication and data collection, objectives, country of study, lockdown dates, sample size, if available, study setting, and study design. We collected data required to answer the review question which were either the reported change in or the numbers of individuals, before and after the start of the pandemic, for each of the study outcomes; TB screening, MDR-TB screening in new and existing patients, TB positivity rate and contact tracing, case notifications, treatment enrolment, diagnostic and treatment delay, clinical diagnosis, presumptive, pulmonary, paediatric, active, latent, and RR/MDR-TB rate, new cases, outpatients, discharged patients and treatment completion, success, and failure. Data on loss to follow-up, re-treated cases, patients not evaluated, sensitive TB, and TB-related deaths before and during the COVID-19 pandemic were also extracted.

### 2.6. Study outcomes

This study had multiple outcomes of interest based on each level of the cascade of care. These outcomes included TB screening, rifampin resistance (RR), multi-drug resistant TB (MDR-TB) screening in new and existing patients, TB positivity rate, diagnostic delay, changes in contact tracing, detection rate, case notifications, treatment delay and clinical diagnosis. Other outcomes were changes in presumptive TB diagnoses, pulmonary, paediatric, active, latent, and sensitive TB, changes in the numbers attending outpatients and discharged inpatients, changes in treatment enrolment, completion, success, and failure, loss to follow-up, re-treated cases, patients not evaluated and TB death. For each study, the percentage change in the number of events was calculated by subtracting the pre and during COVID-19 events and expressing it as a percentage of the pre-COVD19 period. Most studies did not report the populations during the two intervals, so the results could not be standardized.

### 2.7. Assessment of risk of bias of included studies

Two reviewers independently assessed the risk of bias in the included studies. The tool described by Hoy et al. (15) was used to assess the risk of bias of the included studies. They resolved conflicts through consensus, and a third reviewer when consensus was not reached. The tool by Hoy et al. has 10 questions which are used to assess the studies’ external and internal validity. Items 1 to 4 assess a study’s external validity; items 5 to 9 assess internal validity and item 10 assesses biases related to the analysis. An item 11 adds up the scores of the other 10 items to give a summative score that the readers may interpret into low and high-risk categories, but this is subjective, and, in this study, we examined the deficiencies in the individual items rather than an overall score.

### 2.8. Synthesis methods

The characteristics of included studies and the risk of bias were summarized in tables and described narratively in the text. For the main outcomes, we could not conduct the meta-analysis due to the expected differences in the contexts of the included studies, including different lockdown dates and restriction levels, health care system structures, force of COVID-19 infection, policies and pre-existing TB burden and policies. A narrative descriptive synthesis of the percentage change in each of the outcomes was therefore conducted. Findings were summarized using tables and by grouping together similar outcomes across studies. Tableau software [19] was used to create the map of the countries included and the number of studies from each country.

## 3. Results

### 3.1. Study selection

Overall, 7855 records were found from the electronic database and other citation searches, and subsequently 3375 duplicates were removed. Out of the 4480 records, 4343 records were excluded using the title and abstract only. The remaining 135 records were screened using the full text and 109 excluded, resulting in 27 included studies [15,17,20–44] (Figure 1). The reasons for the exclusions were studies that did not include relevant outcomes (n=54), letters to the editor (n=50), qualitative studies (n=2), one was a newsletter and one review (Figure 1).

**Figure 1:**
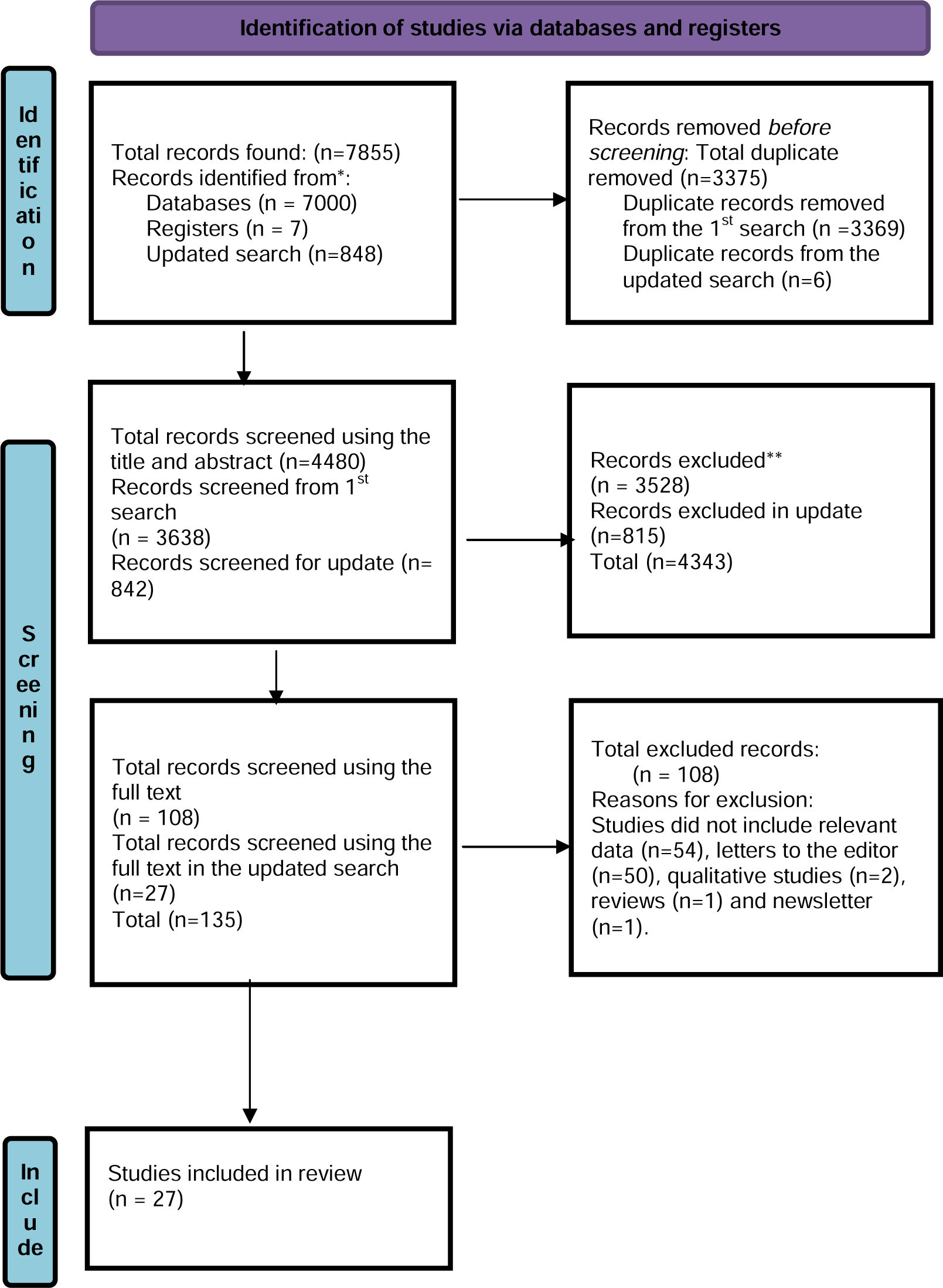
Flowchart of the search and inclusion.

### 3.2. Study characteristics

The included studies were from all regions, as shown on the map in Figure 2. The studies were from the following countries in Africa; Ethiopia, Sierra Leone, Niger, Kenya, Zimbabwe, Malawi, in Asia; Vietnam, India, Singapore, Philippines, China, Iran, Korea, Azerbaijan, South Korea, Israel, Kazakhstan, Kyrgyzstan, Tajikistan, Turkmenistan and Uzbekistan, Australia, South American countries of Brazil and Argentina, and from North America; Canada and Mexico. There were also studies from European countries, namely; Spain, the United Kingdom, Russia, Netherlands, Italy, France, Armenia, Georgia, Portugal, Moldova, Turkey, Ukraine, Albania, Austria, Belarus, Belgium, Bosnia and Herzegovina, Bulgaria, Croatia, Cyprus, Czech Republic, Denmark, Estonia, Finland, Germany, Greece, Hungary, Iceland, Ireland, Italy, Latvia, Lithuania, Montenegro, Norway, Poland, Romania, Serbia, Slovakia, Slovenia, Spain, Sweden, Switzerland, the former Yugoslav and Republic of Macedonia.

**Figure 2:**
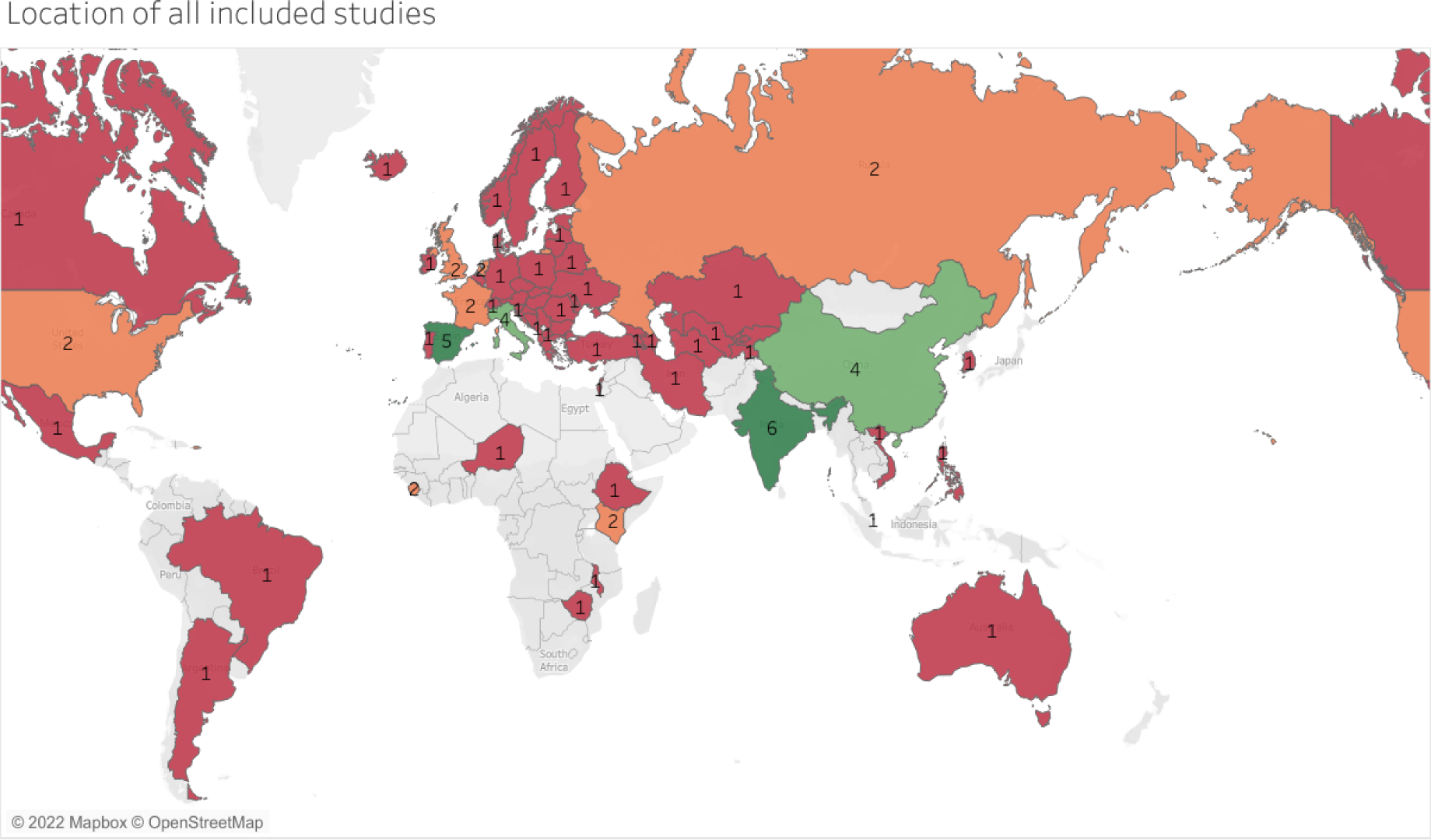
Location of all included studies-the numbers indicate the total number of included studies from each country.

The study designs of the included studies were as follows; one study was described as a surveillance report [45], 12 studies were longitudinal before and after time series [15,24,30–32,34–40], 12 were cohort studies [17,20–22,27–29,33,41–43] and the remaining three were cross-sectional studies [23,25,44]. Ten studies [17,25,26,29,31,32,34,36,37,39] had nationally representative data while others were provincial, and community based. About six studies reported some summary measures of age [30,33–35,38,40], but the other twenty-one studies did not state the age of included participants. The characteristics of the included studies are shown in Supplementary Table 1.

### 3.3. Risk of bias

Twenty-five studies had scores, on the Hoy risk of bias tool, between 6 to 9, suggesting moderate risk of bias and the remaining two studies had moderate scores of 4 and 5, suggesting some high risk of bias. The risk of bias assessment is shown in Supplementary Table 2. Most of the studies scored well on items that measured internal validity with most studies having low risk of selection bias, or information bias. However, some of the studies had deficiencies in external validity. For item 1, nine studies [17,25,26,29,31,32,34,36,39] had a close representation of the country’s national population while it wasn’t clear whether the remaining studies had a close representation of their countries since they were conducted in provinces and local communities. For item 2, the sampling frame in 22 of the 27 studies [15,17,21–23,25–27,29–37,39–43] closely represented the target population. For item 3, only 5 out of the 27 studies [17,31,32,36,37] randomly selected their samples.

### 3.4. Impact of COVID-19 on TB and MDR screening and testing

#### 3.4.1 Changes in TB screening

Six studies, one from Ethiopia [14], one from Vietnam [17], two from India [15,38], and two from China [24,30] investigated changes in screening. Four of these studies [14,15,24,38] reported substantial decreases in TB screening, between 25% and 50% in Ethiopia, India and China, in the COVID-19 period compared to the time before COVID-19. One study from reported a very small decrease of 1% in Vietnam [17] and, conversely another reported a 14% increase in TB screening in Ethiopia [14]. The remaining study, from China, focussed on MDR TB screening and reported a 15.0% decrease in MDR-TB screening in existing patients and a 17% decrease in newly diagnosed TB cases, respectively [30] (Table 1).

**Table 1:** Change in TB and MDR-TB screening, case notification, clinically diagnosed and presumptive TB.

| Study | Country | TB screening | Case notifications | Clinically diagnosed TB | Presumptive TB |
| --- | --- | --- | --- | --- | --- |
| Soko 2021 [37] | Malawi (All provinces) | Not reported | 35.9% reduction in TB notifications in April 2020 compared to the pre-pandemic numbers in April 2016 to March 2020 and April 2020. | Not reported | Not reported |
| Liu 2020[30] | China ( Jiangsu Province) | 15.0% decrease in MDR-TB screening in existing TB cases and a 17.0% decrease in MDR TB between January 2015 to December 2019 and January to May 2020. | 36.5% decrease between January 2015 to December 2019 and January to May 2020. | Not reported | Not reported |
| Srivastava 2021[38] | India (Gurgaon) | 24.9% decrease in TB screening from March 2019 to December 2019 and January 2020 to October 2020. | 15.9% increase between March 2019 and October 2020. | Not reported | Not reported |
| Hazra 2021 [15] | India (South Karnataka) | 49.5% decrease in TB screening between January 2019 and | 49.1% decrease between January 2019 and December 2020. | Not reported | Not reported |
|  |  | December 2020. |  |  |  |
| Geng 2021 [24] | China (Henan province) | 44.5% decrease in TB screening between January to December 2019 and January to December 2020. | Not reported | Not reported | Not reported |
| Arega 2022 [14] | Ethiopia (Addis Ababa) | 14.1% increase in TB screening between April 2019 to March 2020 and April 2020 to March 2021. | 11% decrease between April 2019 to March 2020 and April 2020 to March 2021. | 10.4% decrease between April 2019 to March 2021. | Not reported |
| Hasan 2022 [17] | Vietnam (all provinces) | 1.3% decrease in TB screening between January 2019 to December 2019 and January 2020 to December 2020. | 8.2% decrease between January 2019 to December 2019 and January 2020 to December 2020. | Not reported | Not reported |
| Kwak 2020 [39] | South Korea (all provinces) | Not reported | 28.9% decrease between the first 18 weeks of 2015 to 2019 and the first 18 weeks of 2020. | Not reported | Not reported |
| Thekkur 2021 [42] | Malawi (Lilongwe) | Not reported | 19.1% decrease between March 2019 and February 2020 to March 2020 and February 2021. | 17.1% decrease between March 2019 and February 2020 to March 2020 and February 2021. | 45.6% decrease between March 2019 and February 2020 to March 2020 and February 2021. |
| Dara 2021 [44] | 48 European countries | Not reported | 35.5% decrease between January to June 2019 and January to June 2020. | Not reported | Not reported |
| Lakoh 2021 [20] | Sierra Leone (Free Town) | Not reported | 2.9% decrease between January 2019 to September 2019 and January 2020 to September 2020. | 20.6% decrease between January 2019 to September 2019 and January 2020 to September 2020. | 12.8% decrease between January 2019 to September 2019 and January 2020 to September 2020. |
| Min 2020 [22] | Korea (all provinces) | Not reported | 19.3% decrease between July 2019 to June 2020. | Not reported | Not reported |
| Thekkur 2021 [43] | Zimbabwe (Harare) | Not reported | 33.7% decrease between March 2019-February 2020 to March 2020-February 2021. | 46.0% decrease between March 2019 - February 2020 to March 2020 -February 2021. | 40.6% decrease between March 2019 - February 2020 to March 2020 -February 2021. |
| Fei 2020 [25] | China | Not reported | 24.6% decrease between January - December 2017 to 2019 and January - December 2020. | Not reported | Not reported |
| Feldman 2021 [45] | United States of America (USA) (all States) | Not reported | 19.6% decrease from January - December 2019 and January - December 2020. | Not reported | Not reported |
| Kamakoli 2021 [27] | Iran (Tehran) | Not reported | 32.1% decrease between Feb-June 2016- 2019 to Feb-June 2020. | Not reported | Not reported |
| Arentz 2022 [31] | India (all provinces) | Not reported | 63.3% decrease between January 2017 to April 2021. | Not reported | Not reported |
| Filardo 2022 [32] | USA (US 50 states and the District of Columbia) | Not reported | 8.7% increase between January 2011- December 2011 to January 2021- December 2021. | Not reported | Not reported |
| Golandaj 2021 [36] | India (all provinces) | Not reported | 14.1% decrease between January to September 2019 and January to September 2020. | Not reported | Not reported |
| Mbithi 2021 [41] | Kenya (Nairobi) | Not reported | Not reported | 22.1% decrease between March 2019 to February 2020 and March 2020 to February 2021. | 31.2% decrease between March 2019 to February 2020 and March 2020 to February 2021. |
| Crowder 2021 [46] | Philippines | Not reported | Decline in the daily notification of TB cases by 44.6% (95%CI 38.3%-50.1%) during stable post-quarantine period (day 60 of quarantine) and a plateau in decline from day 60 -174 in comparison to pre-pandemic period (from January to December 2020). | Not reported | Not reported |
| Lungu 2022 [47] | Zambia | Not reported | Monthly decrease in notification 22% (95%CI 19%-24%) in April 2020 in comparison to 2019 (pre-pandemic), After the roll out of the tuberculosis response in July 2020 there was a 45% (95%CI 38%-51%) increase in notification. | Not reported | Not reported |
| Pelissari 2022 [48] | Brazil | Not reported | Decrease from 37.1 in 2019 to 32.6 in 2020 and 34.0 in 2021 per 100000 comparing 2015-2019 notifications and 2020 to 2021 notifications. | Not reported | Not reported |
| Ranasinghe 2022 [49] | 215 countries and focused on 29 high burden countries. | Not reported | Decrease in case notifications in 2020 compared to 2019 in all age groups and region except in African countries. | Not reported | Not reported |

#### 3.4.2 Changes in diagnostic delay and contact tracing

Two studies, each from Italy [40] and India [33] investigated changes in TB diagnostic delay. The two studies [40] [33], reported 35 to 45 days increases in TB diagnostic delay in the COVID-19 period compared to the time before the COVID-19 found in Supplementary Table 3. One study, from Spain [35], reported data on TB contact tracing and showed a 36.1% decrease in TB contact tracing in the COVID-19 period compared to the time before COVID-19 found in Supplementary Table 4.

#### 3.4.3 Changes in detection rate and case notifications

Eighteen studies, two from China [24,30], three from India [15,31,36], two from Malawi [37,42] and USA [26][32] and one each from South Korea, one study that included 48 European countries [44], Sierra Leone [20], Korea [22], Zimbabwe [43], Iran [27],Ethiopia [14],Vietnam and India [38] investigated changes in TB case notification. Sixteen of these studies [14,15,17,20,22,25–27,30,31,36,37,39,42–44] reported decreases between 2.9% in Sierra Leone [20] and 63.3% in India [38] in TB case notifications (Table 1). However, two studies reported increases TB case notifications, of 8.7% in the USA [32] and 15.9% in India [38].

Four studies, each from India [38], Kenya [41], Malawi [42] and Zimbabwe [43], investigated changes in TB positivity rates. Three of these studies reported slight increases, in TB positivity rates in Malawi (4.5%) [42], Kenya (0.1%) [41] and Zimbabwe (2.4%) [43] in the COVID-19 period compared to the time before the COVID-19. In contrast, one study from India [38] reported a 24.9% decrease in TB positivity rate.

One study investigated changes in community and general TB detection rates from Ethiopia [14], and reported 44.7% and 11.8% decreases in these rates, respectively, in the COVID-19 period compared to the time before the COVID-19 found in Supplementary Table 3.

#### 3.4.4 Changes in clinical diagnosis and presumptive TB

Five studies, each from Kenya [41], Malawi [42], Sierra Leone [20], Zimbabwe [43] and Ethiopia [14] reported changes in clinically diagnosed TB. All the studies [14,20,41–43] reported decreases in the number of clinical diagnoses in the COVID-19 period, with the lowest being 10.4% in Ethiopia and the highest being 46.0% in Zimbabwe.

Four studies, each from Kenya [41], Malawi [42], Sierra Leone [20], and Zimbabwe [43] reported changes in presumptive TB. All the studies [20,41–43] reported decreases in the numbers of presumptive TB diagnoses, with the lowest being 12.8% in Sierra Leone [20] and the highest decrease of 45.6% reported from Malawi [42] (Table 1).

#### 3.4.5 Changes in latent, active, pulmonary, and paediatric TB

Two studies, each from India [33] and Spain [35], reported data on changes in pulmonary TB, and reported decreases of 20.0% in India [33] and 50.7% in Spain [35] during the COVID-19 period, compared to the time before COVID-19.

Only one study from India [36] investigated changes in pediatric TB, and reported a 14.1% decrease in pediatric TB cases in the COVID-19 period compared to the before the COVID-19 [36] (Table 2). Two studies, each from Spain [23] and Canada [34] investigated changes in active TB cases and reported decreases of 12.2% and 29.0%, in the two countries, respectively, during the COVID-19 period (Table 2). The same two studies investigated changes in latent TB and reported increases of 30% in Spain [35] and 66.0% in Canada [34] during the COVID-19 period (Table 2).

**Table 2:**
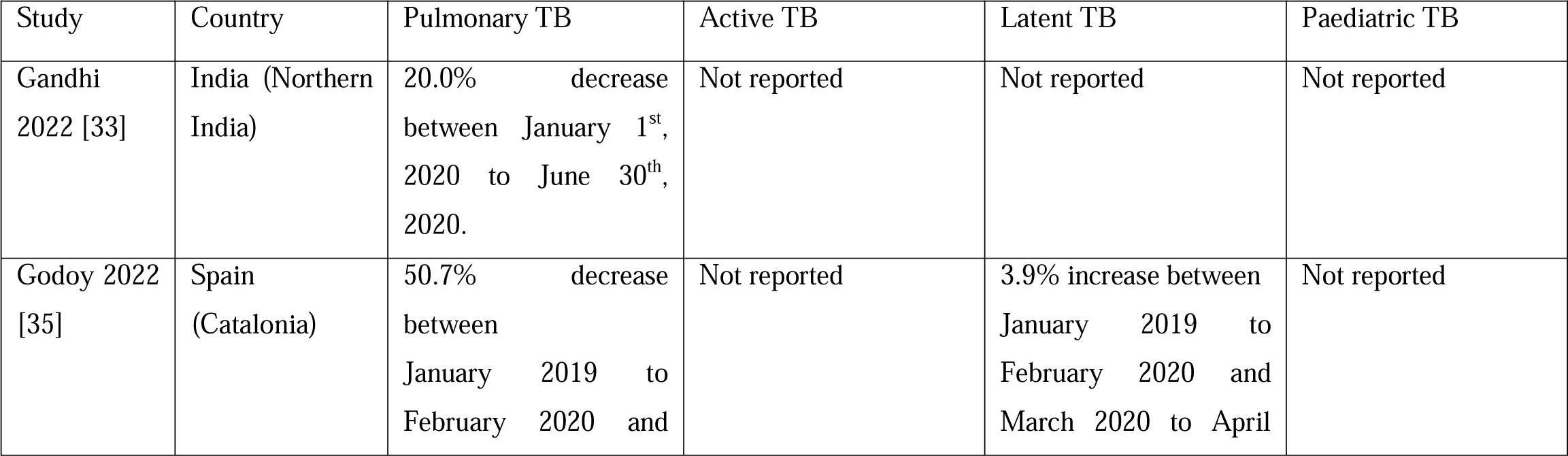
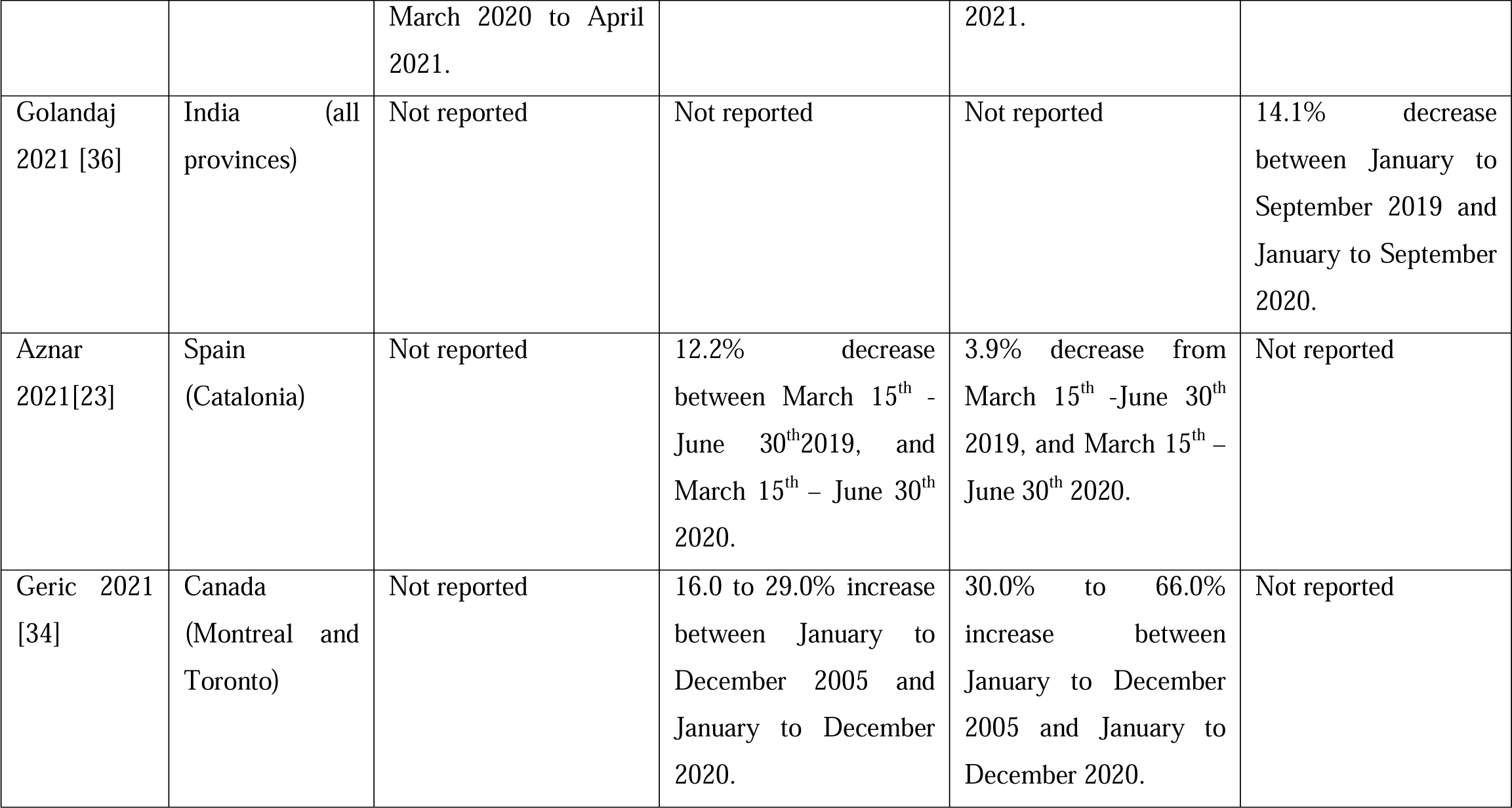
Change in pulmonary, active, latent, and paediatric TB.

| Study | Country | Pulmonary TB | Active TB | Latent TB | Paediatric TB |
| --- | --- | --- | --- | --- | --- |
| Gandhi 2022 [33] | India (Northern India) | 20.0% decrease between January 1<sup>st</sup>, 2020 to June 30<sup>th</sup>, 2020. | Not reported | Not reported | Not reported |
| Godoy 2022 [35] | Spain (Catalonia) | 50.7% decrease between January 2019 to February 2020 and | Not reported | 3.9% increase between January 2019 to February 2020 and March 2020 to April | Not reported |
|  |  | March 2020 to April 2021. |  | 2021. |  |
| Golandaj 2021 [36] | India (all provinces) | Not reported | Not reported | Not reported | 14.1% decrease between January to September 2019 and January to September 2020. |
| Aznar 2021[23] | Spain (Catalonia) | Not reported | 12.2% decrease between March 15<sup>th</sup> - June 30<sup>th</sup>2019, and March 15<sup>th</sup> – June 30<sup>th</sup> 2020. | 3.9% decrease from March 15<sup>th</sup> -June 30<sup>th</sup> 2019, and March 15<sup>th</sup> – June 30<sup>th</sup> 2020. | Not reported |
| Geric 2021 [34] | Canada (Montreal and Toronto) | Not reported | 16.0 to 29.0% increase between January to December 2005 and January to December 2020. | 30.0% to 66.0% increase between January to December 2005 and January to December 2020. | Not reported |

### 3.5 Impact of COVID-19 on TB treatment enrolment and retention

#### 3.5.1 Changes in treatment enrolment and treatment delay

Four studies reported changes in TB treatment enrollment and Rifampin resistant (RR-TB (RR-TB)/MDR-TB treatment enrollment in Kenya [41], Malawi [42], Zimbabwe [43], and 48 European countries [44] (Albania, Armenia, Austria, Azerbaijan, Belarus, Belgium, Bosnia and Herzegovina, Bulgaria, Croatia, Cyprus, Czech Republic, Denmark, Estonia, Finland, France, Georgia, Germany, Greece, Hungary, Iceland, Ireland, Israel, Italy, Kazakhstan, Kyrgyzstan, Latvia, Lithuania, Montenegro, Netherlands, Norway, Poland, Portugal, Republic of Moldova, Romania, Russian Federation, Serbia, Slovakia, Slovenia, Spain, Sweden, Switzerland, Tajikistan, The former Yugoslav, Republic of Macedonia, Turkey, Turkmenistan, Ukraine, United Kingdom and Uzbekistan). All the studies [41–44] reported decreases, between 15.7% in Malawi [42] and 35.0% in Kenya [41] in TB treatment enrollment and RR-TB/MDR-TB treatment enrollment in the COVID-19 period compared to the before the COVID-19 (Table 3). One study from China [30] reported data on treatment completion and showed an 8.0% decrease in treatment completion in the COVID-19 period compared to the time before the COVID-19 (Table 3).

**Table 3:** Change in TB treatment enrolment and completion.

| Study | Country | TB Treatment enrolment | Treatment success rate | Treatment completion |
| --- | --- | --- | --- | --- |
| Mbithi 2021[41] | Kenya (Nairobi) | 35.0% decrease between March 2019 to February 2020 and March 2020 to February 2021. | 2.0% increase between March 2019 to February 2020 and March 2020 to February 2021. | Not reported |
| Thekkur 2021[42] | Malawi (Lilongwe) | 15.7% decrease between March 2019 - February 2020 to March 2020 -February 2021. | 0.1% decrease between March 2019 - February 2020 to March 2020 -February 2021. | Not reported |
| Dara 2021[44] | 48 European countries | 33.5% decrease between April to June 2020. | Not reported | Not reported |
| Thekkur 2021[43] | Zimbabwe (Harare) | 19.1% decrease between March 2019 - February 2020 to March 2020 -February 2021. | 11.6% decrease between March 2019 - February 2020 to March 2020 -February 2021. | Not reported |
| Liu 2020[30] | China (Jiangsu province) | Not reported | Not reported | 8.0% decrease between January 2015 to December 2019 and January to May 2020. |
| Lakoh 2021 [20] | Sierra Leone (Free Town) | Not reported | 15.7% increase between January 2019 to September 2019 and January 2020 to September 2020. | Not reported |
| Min 2020 [22] | Korea (all administrative provinces) | Not reported | 5.9% decrease between July 2019 to June 2020. | Not reported |
| Arega 2022 [14] | Ethiopia (Addis Abba) | Not reported | 17.0% decrease between April 2019 to March 2020 | Not reported |
|  |  |  | and April 2020 to March 2021. |  |
| Hasan 2022 [17] | Vietnam (all provinces) | Not reported | 0.3% decrease between January 2019 to December 2019 and January 2020 to December 2020. | Not reported |

In India, treatment delay increased by 6 days [33] in the COVID-19 period compared to the time before the COVID-19 (Supplementary Table 3).

#### 3.5.2 Changes in numbers of outpatients

Only one study, the multinational study [28], reported data on changes in the numbers of TB outpatients, with data from thirteen countries, which were Australia, Singapore, France, Spain, India, Philippines, Italy, Russia, UK, Mexico, Argentina, Brazil, Niger and Sierra Leone. Four countries Australia, Singapore, France and Spain, from the multinational study [28], reported increases in TB outpatients ranging from 1.0% in Spain up to 40.1% in France (Supplementary Table 4). However, data from nine countries (India, Philippines, Italy, Russia, UK, Mexico, Argentina, Brazil, Niger and Sierra Leone) in the same study [28] showed decreases between 0.5% in Brazil and 71.6% in India (Supplementary Table 4).

#### 3.5.3 Changes in loss to follow-up and patients that were not evaluated

Four studies, each from Kenya [41], Sierra Leone [20], Vietnam (all provinces) [17] and Zimbabwe [43] investigated changes in patients lost to follow-up. Three of these studies [17,20,41] reported decreases in patients lost to follow up, from 0.3% in Kenya [41] and 77.0% in Vietnam [17] in the COVID-19 period compared to the time before the COVID-19 (Table 4). The remaining study [43], reported a slight (0.3%) increase in loss to follow-up in Zimbabwe.

**Table 4.** Change in TB loss to follow-up, failed treatment, re-treated cases and patients not evaluated.

| Study | Country | TB loss to follow-up | Failed treatment | Patients that were not evaluated | Re-treated cases |
| --- | --- | --- | --- | --- | --- |
| Mbithi 2021[41] | Kenya (Nairobi) | 0.3% decrease between March 2019 to February 2020 and March 2020 to February 2021. | 0.3% decrease between March 2019 to February 2020 and March 2020 to February 2021. | 2.2% decrease between March 2019 to February 2020 and March 2020 to February 2021. | Not reported |
| Thekkur 2021 [43] | Zimbabwe (Harare) | 0.3% increase between March 2019-February 2020 to March 2020-February 2021. | 0.2% decrease between March 2019-February 2020 to March 2020-February 2021. | 12.1% decrease between March 2019-February 2020 to March 2020-February 2021. | Not reported |
| Lakoh 2021 [20] | Sierra Leone (Free Town) | 25.4% decrease between January 2019 to September 2019 and January 2020 to September 2020. | 20% decrease between January 2019 to September 2019 and January 2020 to September 2020. | 3.25% increase between January 2019 to September 2019 and January 2020 to September 2020. | Not reported |
| Hasan 2022 [17] | Vietnam (all provinces) | 77% decrease between 2018 and 2020. | 64.2% decrease between January 2019 to December 2019 and January 2020 to December 2020. | 70.8% decrease between January 2019 to December 2019 and January 2020 to December 2020. | Not reported |
| Wang 2021 [21] | China | Not reported | Not reported | Not reported | 76.2% decrease between 2018-2020. |
| Thekkur 2021[42] | Malawi (Lilongwe) | Not reported | Not reported | 0.3% increase between March 2019-February 2020 to March | Not reported |
|  |  |  |  | 2020- February<br>2021. |  |

Five studies, each from Malawi [42], Sierra Leone [20], Zimbabwe [43], Kenya [41] and Vietnam [17], investigated changes in TB patients that were not evaluated. Three of these studies [20,42,43] from Malawi, Sierra Leone and Zimbabwe, reported increases in TB patients that were not evaluated by 0.3% in Malawi [42], 3.25% in Sierra Leone [20] and 12.1% in Zimbabwe [43] in the COVID-19 period compared to the time before the COVID-19. The remaining two studies, reported decreases of 2.2% from Kenya [41] and 70.8% from Vietnam [17] in TB patients that were not evaluated during the COVID-19 period (Table 4).

### 3.6 Impact of COVID-19 on TB treatment outcomes

#### 3.6.1 Changes in TB treatment success rate

Seven studies, each from Malawi [42], Korea [22], Zimbabwe [43], Ethiopia [14], Vietnam [17], Kenya [41] and Sierra Leone [20] investigated changes in TB treatment success rate. Five of these studies, from Malawi, Korea, Zimbabwe, Ethiopia and Vietnam, reported decreases in TB treatment success rate ranging from 0.1% in Malawi [42] to 17.0% in Ethiopia [14], in the COVID-19 period compared to the time before the COVID-19 (Table 3). However, in contrast to the data from those five countries, TB treatment success rate increased by 2.0% and 15.7% in Kenya and Sierra Leone, respectively, in the COVID-19 period compared to the time before the COVID-19.

#### 3.6.2 Changes in discharged inpatients

Two studies, the multinational study [28] from Australia, India, the Philippines, France, Italy, Russia, Spain, UK, Brazil, Singapore, Netherlands and Mexico, and another study from India [38], investigated changes on TB discharged inpatients. The multinational study [28] reported decreases between 6.1% in Philippines and 63.0% in India in discharged patients in the COVID-19 period compared to the time before the COVID-19. The same multinational study [28] also reported increases in discharged TB patients between 12.1% in Singapore and 90.8% in Mexico in the COVID-19 period found in Supplementary Table 5. In India [28], there was an increase in discharged TB patients of 15.4% in in the COVID-19 period (Supplementary Table 4).

#### 3.6.3 Changes in treatment failure and re-treated cases

Four studies, each from Kenya [41], Sierra Leone [20], Zimbabwe [43] and Vietnam [17] investigated changes in TB treatment failure. All the studies [17,20,41,43] reported decreases in the numbers of failed TB treatment, between 0.2% in Zimbabwe [43] and 64.2% in Vietnam [17] (Table 4). The study from China [21] focused on TB retreated cases and reported a 76.2% decrease in re-treated cases in the COVID-19 period compared to the time before the COVID-19 (Table 4).

#### 3.6.5 Changes in drug resistance (DR) occurrence

Five studies, each from India [38], China (25), the 48 European countries [44], Vietnam [17] and Ethiopia [14], investigated changes in RR-TB/MDR-TB occurrence. Three of these studies [17,38,44] reported decreases in RR-TB/MDR-TB of 9.9% in India [38], 1.3% in Vietnam [17] and 33.5% in the 48 European countries [44] (Albania, Armenia, Austria, Azerbaijan, Belarus, Belgium, Bosnia and Herzegovina, Bulgaria, Croatia, Cyprus, Czech Republic, Denmark, Estonia, Finland, France, Georgia, Germany, Greece, Hungary, Iceland, Ireland, Israel, Italy, Kazakhstan, Kyrgyzstan, Latvia, Lithuania, Montenegro, Netherlands, Norway, Poland, Portugal, Republic of Moldova, Romania, Russian Federation, Serbia, Slovakia, Slovenia, Spain, Sweden, Switzerland, Tajikistan, The former Yugoslav, Republic of Macedonia, Turkey, Turkmenistan, Ukraine, United Kingdom and Uzbekistan) in the COVID-19 period (Supplementary Table 5). However, the remaining study, from Ethiopia [14], reported a 27.7% increase in the RR-TB/MDR-TB during the COVID-19 period compared to the time before the COVID-19. The study from China [24] investigated changes in MDR-TB rates and reported a 5.1% increase in the MDR-TB rate during the COVID-19 period compared to the time period before the COVID-19. The study from India [38] reported data on sensitive TB which increased by 12.3% during the COVID-19 period (Supplementary Table 5).

#### 3.6.6 Changes in death due to TB

Five studies, each from India [38], Kenya [41], Malawi [42], Sierra Leone [20], and Vietnam [17] investigated changes in TB deaths. Two of these studies reported slight increases in TB deaths, of 0.8% in Kenya [41] and 2.6% in India [38], in the COVID-19 period compared to the time before the COVID-19. The remaining three studies reported decreases in TB deaths, of 0.6% in Malawi [42], 51.4% in Sierra Leone [20], and 67.0% in Vietnam [17] in the COVID-19 period compared to the time before the COVID-19 (Supplementary Table 5).

## 4. Discussion

This review, which included 27 studies from various countries globally, found that COVID-19 had a significant impact on the cascade of care for TB. The included studies suggested that COVID-19 resulted in substantial decreases in TB screening and diagnosis, as well as decreased treatment enrolment and retention. The findings also suggested that COVID-19 had mixed effects on treatment outcomes, with some studies showing improved outcomes and others showing worse outcomes.

Findings from the included studies suggested that, during COVID-19, TB screening decreased by between 1.3% and 49.5%, MDR TB decreased by between 15% and 17%, clinical TB diagnoses decreased by 10.4% and 46.0%, and case notifications decreased by between 2.9% and 63.3%. Findings from this review could not be compared to other reviews, as there was no other review on the effect of COVID-19 on the TB cascade of care, to the best of our knowledge. It is worth noting that decreases in TB and MDR TB screening could have multiple adverse effects on the health system due to lengthened case detection gap, diagnostic delay, and decreased linkage to care. This may result in increasing TB prevalence, community transmission and incidence. [50–52]. Furthermore, decreases in screening may trigger a resurgence of the disease in countries which were on the road to achieving suppression of the TB. It is therefore important that care is taken, in future health emergencies, to protect key components of the cascade of care of infectious diseases such as TB.

We found that COVID-19 caused changes in some treatment outcomes such as decreased treatment success rates by up to 17%, and increased treatment delays, by up to 6 days, although the impact was less clear on drug resistance rate and death due to TB. Notably, deaths due to TB might may have been attributed to death causes due to the reduction in screening and diagnosis. Likewise, the results of the drug resistance rate might have been reduced due to the restrictions and decrease in screening and diagnosis rates. Again, these findings could not be compared to other reviews since we found no other review on the effects of COVID-19 on TB treatment success, drug resistance and deaths due to TB.

The implications of these findings are significant, as disruptions in the TB care cascade could lead to an upsurge in the number of people living with TB and associated mortality. This could have major consequences for healthcare systems and the economy, particularly in countries where TB is already a major public health concern. One proposition could be to integrate the TB care cascade into universal health coverage as this can be used to manage and identify missing TB patients [55].

However, it’s important to note that this study had several limitations, including the observational nature of the included studies and the presence of confounding variables such as comorbidities and age. Additionally, some of the included studies had small sample sizes, which may have affected the percentage differences reported. The authors could not conduct meta-analysis as anticipated due to the contextual differences of the included studies; thus, a narrative descriptive synthesis was conducted. Another limitation was that many studies did not report population sizes at each point and therefore the analysis could not use standardized results. To address these limitations, larger representative, semi-experimental studies (e.g., interrupted time series), with standardized (by population size) estimates are required in the future research should focus on larger, more representative studies that control for confounding variables and use standardized reporting to facilitate meta-analysis. The strengths of this study include using PRISMA guidelines for its rigorous conduct, and a comprehensive search strategy that was used.

## 5. Conclusion

The pandemic likely had a detrimental impact on the TB care cascade. These findings suggest a need for policies to protect the existing healthcare systems for TB and other communicable, (and, by extension, non-communicable) diseases in future health emergencies. The results of this study must be applied with caution since mostly observational studies, many without standardized population data, were included.

## Funding

This study was not funded.

## Competing interests

All authors declare no conflicts of interest.

## Availability of data, code and other materials

Other study tables, data extraction sheets and details of included studies are attached to this article as study supplementary documents and excel sheets.

## Supporting information

Supplementary Doc

## Data Availability

All data produced in the present work are contained in the manuscript

## Abbreviations

COVID-19: 2019 Coronavirus disease
DR: Drug resistance
MDR: Multidrug resistance
PRISMA: Preferred Reporting Items for Systematic Reviews and Meta-Analysis
TB: Tuberculosis
UK: United Kingdom
USA: United States of America
WHO: World Health Organisation
XDR: Extensively Drug-Resistance
HIV: Hu

## Notes

### Competing Interest Statement

The authors have declared no competing interest.

### Funding Statement

This study did not receive any funding

## References

1. Makam, Parameshwar; Matsa R. “Big Three” Infectious Diseases: Tuberculosis, Malaria and HIV/AIDS [Internet]. 2021. Available from: https://doi.org/10.2174/1568026621666210916170417

2. Glaziou P. Predicted impact of the COVID-19 pandemic on global tuberculosis deaths in 2020. medRxiv [Internet]. 2021;66:2020.04.28.20079582. Available from: doi: https://doi.org/10.1101/2020.04.28.20079582

3. Thomas Lynch. Global Pandemic | TB Alliance [Internet]. TB Alliance. 2022. Available from: https://www.tballiance.org/why-new-tb-drugs/global-pandemic

4. World Health Organization W. Global Tuberculosis Programme [Internet]. Glob. TB Rep. 2021. Available from: https://www.who.int/teams/global-tuberculosis-programme/the-end-tb-strategy

5. Badalov E, Blackler L, Scharf AE, Matsoukas K, Chawla S, Voigt LP, et al. COVID-19 double jeopardy: the overwhelming impact of the social determinants of health. Int J Equity Health [Internet]. BioMed Central; 2022;21:1–8. Available from: https://doi.org/10.1186/s12939-022-01629-0

6. Desson, Z., Weller, E., McMeekin, P., & Ammi M. An analysis of the policy responses to the COVID-19 pandemic in France, Belgium, and Canada. Heal policy Technol [Internet]. 2020;9(4), 430–. Available from: https://doi.org/10.1016/j.hlpt.2020.09.002

7. CDC. Treating TB During the Time of COVID-19 [Internet]. Glob. Heal. 2021. Available from: https://www.cdc.gov/globalhealth/stories/2020/tb-covid.html

8. Okereke, M., Ukor, N. A., Adebisi, Y. A., Ogunkola, I. O., Favour Iyagbaye, E., Adiela Owhor, G., & Lucero-Prisno, D. E. 3rd. Impact of COVID-19 on access to healthcare in low- and middle-income countries: Current evidence and future recommendations. Int J Health Plann Manage [Internet]. 2021;36(1), 13–. Available from: https://doi.org/10.1002/hpm.3067

9. Yasobant S, Bhavsar P, Kalpana P, Memon F, Trivedi P, Saxena D. Contributing factors in the tuberculosis care cascade in india: A systematic literature review. Risk Manag Healthc Policy [Internet]. 2021;14:3275–86. Available from: https://doi.org/10.2147/RMHP.S322143

10. Subbaraman R, Nathavitharana RR, Satyanarayana S, Pai M, Thomas BE, Chadha VK, et al. The Tuberculosis Cascade of Care in India’s Public Sector: A Systematic Review and Meta-analysis. PLoS Med. 2016;13:1–38.

11. Subbaraman, R., Nathavitharana, R. R., Mayer, K. H., Satyanarayana, S., Chadha, V. K., Arinaminpathy, N., & Pai M. Constructing care cascades for active tuberculosis: A strategy for program monitoring and identifying gaps in quality of care. PLoS Med. 2019;16(2), e10.

12. Mukherjee, A., & Parashar R. Impact of the COVID-19 pandemic on the human resources for health in India and key policy areas to build a resilient health workforce. Gates open Res [Internet]. 2020;4, 159. Available from: https://doi.org/10.12688/gatesopenres.13196.1

13. Zhou S, Van Staden Q, Toska E. Resource reprioritisation amid competing health risks for TB and COVID-19. Int J Tuberc Lung Dis [Internet]. 2020;24:1215–6. Available from: http://dx.doi.org/10.5588/ijtld.20.0

14. Arega B, Negesso A, Taye B, Weldeyohhans G, Bewket B, Negussie T, et al. Impact of COVID-19 pandemic on TB prevention and care in Addis Ababa, Ethiopia: A retrospective database study. BMJ Open [Internet]. 2022;12. Available from: doi:10.1136/bmjopen-2021-053290

15. Hazra D, Chawla K, Shenoy VP, Pandey AK, Nayana S. Journal of Infection and Public Health The aftermath of COVID-19 pandemic on the diagnosis of TB at a tertiary care hospital in India. J Infect Public Health [Internet]. King Saud Bin Abdulaziz University for Health Sciences; 2021;14:1095–8. Available from: https://doi.org/10.1016/j.jiph.2021.07.001

16. WHO. WHO Advice for the public [Internet]. 2020. Available from: https://www.who.int/emergencies/diseases/novel-coronavirus-2019/advice-for-public

17. Hasan T, Nguyen VN, Nguyen HB, Nguyen TA, Le HTT, Pham CD, et al. Retrospective Cohort Study of Effects of the COVID-19 Pandemic on Tuberculosis Notifications, Vietnam, 2020. Emerg Infect Dis [Internet]. 2022;28:684–92. Available from: doi:10.3201/eid2803.211919

18. Page MJ, Moher D, Bossuyt PM, Boutron I, Hoffmann TC, Mulrow CD, et al. PRISMA 2020 explanation and elaboration: Updated guidance and exemplars for reporting systematic reviews. BMJ [Internet]. 2021;372. Available from: doi: https://doi.org/10.1136/bmj.n160

19. Tableau. Mission. Tableau (version 9.1) [Internet]. 2015. Available from: http://mission.tableau.com/#/mission/%3E. [Google Scholar]

20. Lakoh S, Jiba DF, Baldeh M, Adekanmbi O, Barrie U, Seisay AL, et al. Impact of covid-19 on tuberculosis case detection and treatment outcomes in sierra leone. Trop Med Infect Dis [Internet]. 2021;6. Available from: doi: 10.3390/TROPICALMED6030154

21. Wang X, He W, Lei J, Liu G, Huang F, Zhao Y. Impact of COVID-19 Pandemic on Pre-Treatment Delays, Detection, and Clinical Characteristics of Tuberculosis Patients in Ningxia Hui Autonomous Region, China. Front Public Heal [Internet]. 2021;9:1–8. Available from: doi:10.3389/fpubh.2021.644536

22. Min J, Kim HW, Koo HK, Ko Y, Oh JY, Kim J, et al. Impact of COVID-19 Pandemic on the National PPM Tuberculosis Control Project in Korea: the Korean PPM Monitoring Database between July 2019 and June 2020. J Korean Med Sci [Internet]. 2020;35:e388. Available from: doi:10.3346/jkms.2020.35.e388

23. Aznar ML, Espinosa-Pereiro J, Saborit N, Jové N, Sánchez Martinez F, Pérez-Recio S, et al. Impact of the COVID-19 pandemic on tuberculosis management in Spain. Int J Infect Dis [Internet]. 2021;108:300–5. Available from: doi:10.1016/j.ijid.2021.04.075

24. Geng Y, Li G, Zhang L. The Impact of COVID-19 Interventions on Influenza and Mycobacterium Tuberculosis Infection. Front Public Heal [Internet]. 2021;9:1–6. Available from: doi:10.3389/fpubh.2021.672568

25. Fei H, Yinyin X, Hui C, Ni W, Xin D, Wei C, et al. The impact of the COVID-19 epidemic on tuberculosis control in China. Lancet Reg Heal - West Pacific [Internet]. Elsevier Ltd; 2020;3:100032. Available from: https://doi.org/10.1016/j.lanwpc.2020.100032

26. Deutsch-Feldman M, Pratt RH, Price SF, Tsang CA SJ. Tuberculosis — United States, 2020 MMWR. 2020.

27. Mansour Kargarpour Kamakoli, Shima Hadifar, Sharareh Khanipour, Shayan Mostafaei, Seyed Davar Siadat, Farzam Vaziria, Ghazaleh Farmanfarmaei AF. Tuberculosis under the Infl uence of COVID-19 Lockdowns: Lessons from Tehran, Iran. mSphere [Internet]. 2021;2019:2019–22. Available from: doi:10.1128/mSphere.00076-21

28. Migliori GB, Thong PM, Akkerman O, Alffenaar JW, Álvarez-Navascués F, Assao-Neino MM, et al. Worldwide Effects of Coronavirus Disease Pandemic on Tuberculosis Services, January–April 2020. Emerg Infect Dis [Internet]. 2020;26:2709–12. Available from: doi:10.3201/eid2611.203163

29. Arega B, Negesso A, Taye B, Weldeyohhans G, Bewket B, Negussie T, et al. Impact of COVID-19 pandemic on TB prevention and care in Addis Ababa, Ethiopia: A retrospective database study. BMJ Open. 2022;12:1–6.

30. Qiao Liu, Peng Lu, Ye Shen, Changwei Li, Jianming Wang, Limei Zhu, Wei Lu LM. Collateral Impact of the Covid-19 Pandemic on Tuberculosis Control in Jiangsu Province, China Qiao. Oxford Univ Press Infect Dis Soc Am [Internet]. 2020;0:1–14. Available from: http://dx.doi.org/10.1080/07853890.2020.1840620

31. Arentz M, Ma J, Zheng P, Vos T, Murray CJL, Kyu HH. The impact of the COVID-19 pandemic and associated suppression measures on the burden of tuberculosis in India. BMC Infect Dis [Internet]. BioMed Central; 2022;22:1–8. Available from: https://doi.org/10.1186/s12879-022-07078-y

32. Filardo TD, Feng P, Pratt RH, Price SF, Self JL. Tuberculosis — United States, 2021. MMWR Morb Mortal Wkly Rep 2022 Mar 25;71(12)441-446 [Internet]. 2022;71. Available from: doi: 10.15585/mmwr.mm7112a1

33. Gandhi AP, Kathirvel S, Rehman T. Effect of COVID-19 lockdown on the pathway of care and treatment outcome among patients with tuberculosis in a rural part of northern India: a community-based study. J Rural Med [Internet]. 2022;17:59–66. Available from: doi:10.2185/jrm.2021-039

34. Geric C, Saroufim M, Landsman D, Richard J, Benedetti A, Batt J, et al. Impact of COVID-19 on Tuberculosis Prevention and Treatment in Canada: A Multicenter Analysis of 10 833 Patients. J Infect Dis [Internet]. 2022;225:1317–20. Available from: doi:10.1093/infdis/jiab608

35. Godoy P, Parrón I, Barrabeig I, Caylà JA, Clotet L, Follia N, et al. Impact of the COVID-19 pandemic on contact tracing of patients with pulmonary tuberculosis. Eur J Public Health [Internet]. 2022;1–5. Available from: doi:10.1093/eurpub/ckac031

36. Golandaj JA. Since January 2020 Elsevier has created a COVID-19 resource centre with free information in English and Mandarin on the novel coronavirus COVID-19. The COVID-19 resource centre is hosted on Elsevier Connect, the company’s public news and information. Glob Ecol Conserv [Internet]. 2021; Available from: doi:10.1016/j.gecco.2022.e02270

37. Soko RN, Burke RM, Feasey HRA, Sibande W, Nliwasa M, Henrion MYR, et al. Effects of coronavirus disease pandemic on tuberculosis notifications, malawi. Emerg Infect Dis [Internet]. 2021;27:1831–9. Available from: https://doi.org/10.3201/eid2707.210557

38. Srivastava S, Jaggi N. ScienceDirect Original article TB positive cases go up in ongoing COVID-19 pandemic despite lower testing of TBLJ: An observational study from a hospital from. Indian J Tuberc [Internet]. Elsevier Ltd; 2021; Available from: https://doi.org/10.1016/j.ijtb.2021.04.014

39. Kwak N, Hwang SS, Yima AJ. Effect of COVID-19 on Tuberculosis Notification, South Korea. Emerg Infect Dis [Internet]. 2020;26:2506–8. Available from: doi:10.3201/EID2610.202782

40. Di Gennaro F, Gualano G, Timelli L, Vittozzi P, Di Bari V, Libertone R, et al. Increase in tuberculosis diagnostic delay during first wave of the covidLJ19 pandemic: Data from an Italian infectious disease referral hospital. Antibiotics [Internet]. 2021;10:1–10. Available from: doi: 10.3390/antibiotics10030272

41. Irene Mbithi,† PT, Jeremiah Muhwa Chakaya, Elizabeth Onyango, Philip Owiti, Ngugi Catherine Njeri AMVK, Satyanarayana S, Shewade HD, Mohammed Khogali, Rony Zachariah, I. D. Rusen SDB and ADH. Assessing the Real-Time Impact of COVID-19 on TB and HIV Services: The Experience and Response from Selected Health Facilities in Nairobi, Kenya. Trop Med Infect Dis [Internet]. 2021;6. Available from: doi:10.3390/tropicalmed6020094

42. Thekkur P, Tweya H, Phiri S, Mpunga J, Kalua T, Kumar AM V, et al. Assessing the Impact of COVID-19 on TB and HIV Programme Services in Selected Health Facilities in Lilongwe, Malawi: Operational Research in Real Time. Trop Med Infect Dis [Internet]. 2021;1211. Available from: https://doi.org/10.3390/tropicalmed

43. Thekkur P, Takarinda KC, Timire C, Sandy C, Apollo T, Kumar AMV, et al. Operational research to assess the real-time impact of covid-19 on tb and hiv services: The experience and response from health facilities in Harare, Zimbabwe. Trop Med Infect Dis [Internet]. 2021;6. Available from: doi: 10.3390/tropicalmed6020094

44. Dara M, Kuchukhidze G, Yedilbayev A, Perehinets I, Schmidt T, Van Grinsven WL, et al. Early COVID-19 pandemic’s toll on tuberculosis services, WHO European Region, January to June 2020. Eurosurveillance [Internet]. 2021;26:27–35. Available from: doi: 10.2807/1560-7917.ES.2021.26.24.2100231

45. Deutsch-feldman M, Pratt RH, Price SF, Tsang CA, Self JL. Tuberculosis — United States, 2020. MMWR Morb Mortal Wkly Rep 2021 Mar 26;70(12)409-414 [Internet]. 2021;70. Available from: doi: 10.15585/mmwr.mm7012a1 PMID: 33764959; PMCID: PMC7993554.

46. Crowder R, Geocaniga-Gaviola DM, Fabella RA, Lim A, Lopez E, Kadota JL, et al. Impact of shelter-in-place orders on TB case notifications and mortality in the Philippines during the COVID-19 pandemic. J Clin Tuberc Other Mycobact Dis [Internet]. Elsevier Ltd; 2021;25:100282. Available from: https://doi.org/10.1016/j.jctube.2021.100282

47. Lungu PS, Kerkhoff AD, Muyoyeta M, Kasapo CC, Nyangu S, Kagujje M, et al. Interrupted time-series analysis of active case-finding for tuberculosis during the COVID-19 pandemic, Zambia. Bull World Health Organ. 2022;100:205–15.

48. Daniele M Pelissari, Patricia Bartholomay, Fernanda Dockhorn Costa Johansen FAD-Q. Impact of the Covid-19 Pandemic on the Furniture Market in Serbia. 15th Int Sci Conf WoodEMA 2022 - Cris Manag Saf Foresight For Sect SMES Oper Glob Environ. 2022;13:221–6.

49. Ranasinghe L, Achar J, Gröschel MI, Whittaker E, Dodd PJ, Seddon JA. Global impact of COVID-19 on childhood tuberculosis: an analysis of notification data. Lancet Glob Heal. 2022;10:e1774–81.

50. Telisinghe L, Shaweno D, Hayes RJ, Dodd PJ AH. The effect of systematic screening of the general population on TB case notification rates. Int J Tuberc Lung Dis. 2021;2021 Dec 1.

51. World Health Organization. WHO consolidated guidelines on tuberculosis. Modul 2 373 Syst Screen Tuberc Dis 2021 [Internet]. 2021; Available from: https://apps.who.int/iris/bitstream/handle/10665/340255/9789240022676-eng.pdf. 375

52. Lonnroth K, Corbett E, Golub J, Godfrey-Faussett P, Uplekar M, Weil D et al. 376. Systematic screening for active tuberculosis: rationale, definitions and key considerations. 377. Int J Tuberc Lung Dis 2013;17(3)289-98 378. 2013;

53. Tedla K, Medhin G, Berhe G, Mulugeta A, Berhe N. Delay in treatment initiation and its association with clinical severity and infectiousness among new adult pulmonary tuberculosis patients in Tigray, northern Ethiopia. BMC Infect Dis [Internet]. BMC Infectious Diseases; 2020;20:1–10. Available from: https://doi.org/10.1186/s12879-020-05191-4

54. Tyagi SK and R. The impact of COVID-19 on tuberculosis: challenges and opportunities. Ther Adv Infect Diseas [Internet]. 2021;9:259–61. Available from: doi: 10.1177/20499361211016973

55. Padayatchi N, Daftary A, Naidu N, Naidoo K, Pai M. Tuberculosis: Treatment failure, or failure to treat? Lessons from India and South Africa. BMJ Glob Heal [Internet]. 2019;4:1–6. Available from: doi:10.1136/%0Abmjgh-2018-001097

