## Supplementary Table for "Impact of COVID-19 on the cascade of care for tuberculosis: A systematic review"

**Supplementary tables**

**Supplementary Table 1- Characteristics of the included studies**

| Study | Study design | Country | Setting | Outcomes | Lockdown period | Number of participants | Study period | Findings |
|---|---|---|---|---|---|---|---|---|
| Crowder 2021 | Time series analysis | Philippines | Hospitals (Urban) | TB case notification | | Not captured | 1st January to 31st December 2020 | Decline in the daily notification of TB cases by 44.6% (95%CI 38.3%-50.1%) during stable post-quarantine period (day 60 of quarantine) and a plateau in decline from day 60 -174 in comparison to pre-pandemic period (from January to December 2020). |
| Lungu 2022 | Time series analysis | Zambia | Hospitals (Urban and rural) | TB case notification | | Not captured | January 2019 to September 2021 | Monthly decrease in notification 22% (95%CI 19%-24%) in April 2020 in comparison to 2019 (pre-pandemic), After the roll out of the tuberculosis response in July 2020 there was a 45% (95%CI 38%-51%) increase in notification. |
| Pelissari 2022 | Surveillance report | Brazil | National Information System (Urban and rural) | TB case notification | | Not captured | 2015-2021 | Decrease from 37.1 in 2019 to 32.6 in 2020 and 34.0 in 2021 per 100000 comparing 2015-2019 notifications and 2020 to 2021 notifications. |
| Ranasinghe 2022 | Time series | 215 countries and focused on 29 high burden countries (Papua New Guinea, Indonesia, DR Congo, Philippines, India, | WHO notification | TB case notification | Varied between countries | Not captured | 2014-2020 | Decrease in case notifications in 2020 compared to 2019 in all age groups and region except in African countries. |

| | | | | | | | | |
|---|---|---|---|---|---|---|---|---|
| | | Angola, Pakistan, Sierra Leone, Myanmar, Viet Nam, Bangladesh, Nigeria, Mozambique, Congo (Brazzaville), Kenya, Uganda, Brazil, Gabon, Central African Republic, China, Lesotho, Namibia, Tanzania, North Korea, Thailand, Mongolia, Zambia, Ethiopia and South Africa). | | | | | | |
| Liu 2020 | Longitudinal (before and after) time series study | China (Jiangsu Province) | Hospital (Urban and rural) | TB case notification, MDR TB screening and treatment completion. | January 23rd, 2020 | 143,250 | January 2015 to December 2019 and January to May 2020. | Our analysis suggests a substantial reduction between 36%–52% in tuberculosis notifications in 2020 compared to 2015–2019. |
| Srivastava 2021 | Longitudinal (before and after) time series study | India (Gurgaon) | Hospital (Urban and rural) | TB diagnosis, case notifications, positivity rate, RR and sensitive TB rate, discharge inpatients and deaths. | March 24th, 2020 | For testing samples: 484 in 2020, 644 in 2019 For notification samples 146 in | March 2019 to December 2019 and January 2020 to October 2020. | Our study reported an increase in confirmed TB cases in 2020 as compared to 2019. |

| | | | | | | | | |
|---|---|---|---|---|---|---|---|---|
| | | | | | | 2020, 127 in 2019 | | |
| Hazra 2021 | Longitudinal (before and after) time series study | India (South Karnataka) | Hospital (Rural) | TB diagnosis and case notifications. | March 24th, 2020 | Not Stated | January 2019 to December 2020. | Our study reported a significant decrease in TB diagnosis and active TB case detection. |
| Kwak 2020 | Longitudinal (before and after) time series study | South Korea (all provinces) | Community (Urban and rural) | TB case notification | February 23rd, 2020 | Not Stated | First 18 weeks of 2015 to 2019 and the first 18 weeks of 2020. | Our study reported a significant decrease in TB diagnosis and notification as the surge of COVID-19 infection in South Korea. |
| Gennaro 2021 | Longitudinal (before and after) time series study | Italy (Rome) | Hospital (Urban) | TB diagnostic and treatment delay. | March 10$^{th}$, 2020 | 201 patients in 2019 115patients in 2020 | March 2019 to August 2020. | Our study reported higher TB diagnostic delay, a reduction in hospitalization and greater severity of clinical presentations during the COVID-19 pandemic. |
| Mbithi 2021 | Cohort study | Kenya (Nairobi) | Hospital (Urban) | TB treatment enrolment, failure and success rate, loss to follow-up, patients not evaluated and deaths. | March 20$^{th}$, 2020 | Not Stated | March 2019 to February 2020 and March 2020 to February 2021. | Our study reported that the programmatic interventions implemented during the COVID-19 period were associated with improved case detection and treatment outcomes during the COVID-19 period, suggesting that monthly real-time surveillance is useful during unprecedented events. |
| Thekkur 2021 | Cohort study | Malawi (Lilongwe) | Hospital (Urban and rural) | TB case notification, treatment enrolment, clinical diagnosis, positivity rate, presumptive TB, treatment success and failure rate, patients that were not evaluated and deaths. | April 18$^{th}$, 2020 17 January 2021 | Not Stated | March 2019 and February 2020 to March 2020 and February 2021. | Our study reported a decline in TB case detection and treatment outcomes for TB during the COVID-19 pandemic. |

| | | | | | | | | |
|---|---|---|---|---|---|---|---|---|
| Dara 2021 | Cross-sectional survey | 48 European countries | Community (Urban and rural) | TB case notifications, TB treatment | The lockdown period varied in countries. | Not stated | January to June 2019 and January to June 2020. | Our study reported a substantial decrease in TB notifications in Q2 2020 in the WHO European Region. This delay or lack of diagnosis can lead to ongoing transmission of the disease to close contacts, increased severity of TB disease and a potential increase in case fatality. |
| Lakoh 2021 | Cohort study (Retrospective) | Sierra Leone (Free Town) | Hospital (Urban) | TB notification TB treatment | April 1st, 2020 | Not Stated | January 2019 to September 2019 and January 2020 to September 2020. | Our study reported COVID-19 negative impacts on TB care at the largest treatment centre in Sierra Leone. |
| Wang 2021 | Cohort study | China (Ningxia Hui) | Hospital (Urban) | TB case notification TB treatment TB Patient delay | January 23rd, 2020 | Not stated | 2018-2020 | Our study reported a reduction in cases of TB notification in Ningxia due to the COVID-19 pandemic. |
| Min 2020 | Cohort study | Korea (all provinces) | Hospital (Urban) | TB case notification TB treatment success rate | February 23rd, 2020 | Not stated | July 2019 to June 2020 | Our study reported the COVID-19 pandemic's enormous potential to hinder the efforts of TB services in prevention, case detection, and management, particularly in resource-limited settings. |
| Aznar 2021 | Cross-sectional survey | Spain (all provinces) | Hospital (Urban) | TB case notification | March 14th, 2020 | Not stated | March 15-June 30, 2019 and March 15- June 30, 2020. | Our study reported an increase in LTBI infection and active TB in children whose household had contact with patients. This reflects increased household transmission due to the anti-COVID-19 measures. |
| Thekkur 2021 | Cohort study | Zimbabwe (Harare) | Hospital (Urban) | TB case notification, treatment enrolment, clinical diagnosis, positivity rate, presumptive | March 30th, 2020 | Not stated | March 2019 and February 2020 to March 2020 and February 2021. | Our study reported a declining trend in TB case detection and treatment outcomes. |

| | | | | | | | | |
|---|---|---|---|---|---|---|---|---|
| | | | | TB, treatment success and failure rate and patients that were not evaluated. | | | | |
| Geng 2021 | Longitudinal (before and after) time series study | China (Henan province) | Hospital (Urban and rural) | TB diagnosis and MTB cumulative rate. | January 23rd, 2020 | Not stated | January to December 2019 and January to December 2020. | Our study reported less effect of a non-pharmaceutical public health intervention on MTB transmission in 2020. |
| Fei 2020 | Cross-sectional survey | China (all provinces) | Hospital (Urban and rural) | TB case notifications | January 23rd, 2020 | Not stated | January – December 2017 to 2019 and January - December 2020 | Our study reported reductions in TB notifications and follow-up examinations in China during the COVID-19 pandemic and this may cause an upsurge in TB cases in the nearest future. |
| Feldman 2021 | Surveillance report | USA (all states) | Community (Urban and rural) | TB case notifications | Days between March 19th to April 7th, 2020 (it varies with the area). | Not stated | January - December 2019 and January - December 2020. | Our study reported incidence a 20% TB incidence decrease in 2020 as compared to 2019 cases. |
| Kamakoli 2021 | Longitudinal (before and after) time series study | Iran (Tehran) | Research Institute (Urban and rural) | TB case notifications | March 13th, 2020 | Not stated | February -June 2016-2019 to February -June 2020 | Our study reported a significant decrease in TB case identification in Tehran, in 2020. |
| Migliori 2020 | Cohort study | 16 countries | Hospital (Urban) | TB new cases discharged inpatients and outpatients. | The lockdown period varied in countries. | Not Stated | January-April 2019 and January-April 2020. | Our study reported reductions in TB-related hospital discharges, newly diagnosed cases of active TB, total active TB outpatient visits and new LTBI and LTBI outpatient visits in the first 4 months of 2020. |
| Arega 2022 | Cohort study | Ethiopia (Addis Ababa) | Research Institute (Urban) | TB screening, case notification, detection | April 8th, 2020 | Not Stated | April 2019 to March 2020 and April | Our study reported a negative impact of the COVID-19 pandemic on TB service |

| | | | | | | | | |
|---|---|---|---|---|---|---|---|---|
| | | | | rate, clinical diagnosis, treatment success and MDR-RR rate. | | | 2020 to March 2021. | indicators in Addis Ababa, Ethiopia. |
| Arentz 2022 | Longitudinal (before and after) time series study | India (all provinces) | Research Institute (Urban and rural) | TB case notification | March $24^{th,}$ 2020 | Not Stated | January 2017 to April 2021. | Our study a large difference between reported TB cases in India and those expected in the absence of the pandemic. |
| Filardo 2022 | Longitudinal (before and after) time series study | USA (US 50 states and the District of Columbia) | Community (Urban and rural) | TB case notification | March $19^{th}$ to April $7^{th}$ 2020 | Not Stated | January 2011- December 2011 to January 2021- December 2021. | Our study reported a significant decrease in TB case notifications in the USA. |
| Gandhi 2022 | Retrospective cohort study | India (Northern India) | Community (Rural) | TB diagnostic delay, treatment delay and pulmonary TB. | March $24^{th}$, 2020 | 103 | January $1^{st}$, 2020 to June $30^{th}$, 2020. | Our study reported a significant decrease in pulmonary TB notification and an increase in diagnostic delay in Northern India. |
| Geric 2021 | Longitudinal (before and after) time series study | Canada (Montreal and Toronto) | Hospital (Urban) | Active and latent TB. | March 14 and 17 2020 in the Quebec and Ontario provinces respectively. | 10833 | January to December 2005 and January to December 2020 | Our study reported a significant decrease in active and latent TB treatment in Ontario and Quebec provinces. The enactment of public health emergency measures against COVID-19 in Canada weakened the measures for tuberculosis control and treatment. |
| Godoy 2022 | Longitudinal (before and after) time series study | Spain (Catalonia) | 1 Hospital in Catalonia (Northern Spain) (Urban) | TB contact tracing, pulmonary and latent TB. | March $14^{th}$, 2020 | 6363 | January 2019 to February 2020 and March 2020 to April 2021. | Our study reported less exhaustive TB and LTBI case detection, though an increase in LTBI was observed during the pandemic. |
| Golandaj 2021 | Longitudinal (before and after) time series study | India (all provinces) | Research Institute (Urban and rural). | Case notifications and paediatric TB. | March $24^{th}$, 2020 | Not Stated | January to September 2019 and 2020. | Our study reported a significant decrease in paediatric TB during the COVID-19 pandemic. |

| | | | | | | | | |
|---|---|---|---|---|---|---|---|---|
| Hasan 2022 | Retrospective cohort study | Vietnam (all provinces) | Vietnam's 63 provinces (Community, Urban and rural) | TB screening, notification, treatment success and failure rate, loss to follow-up, patients that were not evaluated, RR MDRTB and deaths. | April 1st, 2020 | Not Stated | January 2019 to December 2019 and January 2020 to December 2020. | Our study reported a limited decrease in TB notifications in Vietnam during the first year of the COVID-19 pandemic. |
| Soko 2020 | Interrupted time series study | Malawi | Hospital (Urban and rural) | TB notifications | April 18th, 2020 17 January 2021 | Not stated | April 2016 to March 2020 and April 2020. | Our study reported a 35.9% reduction in TB notifications in April 2020 as compared to the pre-pandemic numbers from April 2016 to March 2020 and April 2020. |

**Supplementary Table 2: Hoy risk of bias assessment**

| Studies | I | II | III | IV | V | VI | VII | VIII | IX | X | Summary of the overall Risk of bias for each study (XI) |
|---|---|---|---|---|---|---|---|---|---|---|---|
| Liu 2021 | 0 | 1 | 0 | 1 | 1 | 1 | 1 | 0 | 1 | 1 | 7 |
| Srivastava 2021 | 0 | 0 | 0 | 0 | 1 | 1 | 1 | 1 | 1 | 1 | 6 |
| Hazra 2021 | 0 | 1 | 0 | 1 | 1 | 1 | 1 | 1 | 1 | 1 | 8 |
| Kwak 2020 | 1 | 1 | 0 | 1 | 1 | 0 | 1 | 1 | 1 | 1 | 8 |
| Gennaro 2021 | 0 | 1 | 0 | 0 | 1 | 0 | 1 | 1 | 1 | 1 | 6 |
| Mbithi 2021 | 0 | 1 | 0 | 1 | 1 | 1 | 1 | 1 | 1 | 1 | 8 |
| Thekkur 2021 Malawi | 0 | 1 | 0 | 1 | 1 | 1 | 1 | 1 | 1 | 1 | 8 |
| Dara 2020 | 0 | 0 | 0 | 1 | 1 | 1 | 1 | 1 | 1 | 1 | 7 |
| Lakoh 2021 | 0 | 0 | 0 | 1 | 0 | 1 | 1 | 1 | 1 | 1 | 6 |
| Wang 2021 | 0 | 1 | 0 | 1 | 0 | 1 | 1 | 1 | 1 | 1 | 7 |
| Min 2020 | 0 | 1 | 0 | 0 | 0 | 0 | 1 | 1 | 1 | 1 | 5 |
| Aznar 2021 | 0 | 1 | 0 | 0 | 1 | 1 | 0 | 1 | 1 | 1 | 6 |
| Thekkur 2021 Zimbabwe | 0 | 1 | 0 | 1 | 1 | 1 | 1 | 1 | 1 | 1 | 8 |
| Geng 2021 | 0 | 0 | 0 | 1 | 0 | 1 | 1 | 1 | 1 | 1 | 6 |
| Fei 2020 | 1 | 1 | 0 | 0 | 1 | 1 | 1 | 1 | 1 | 1 | 8 |
| Feldman 2020 | 1 | 1 | 0 | 0 | 1 | 0 | 1 | 1 | 1 | 1 | 7 |

| | | | | | | | | | | |
|---|---|---|---|---|---|---|---|---|---|---|
| Kamakoli 2021 | 0 | 1 | 0 | 1 | 1 | 0 | 1 | 1 | 1 | 1 | 7 |
| Migliori 2020 | 0 | 0 | 0 | 0 | 0 | 0 | 1 | 1 | 1 | 1 | 4 |
| Arega 2022 | 1 | 1 | 0 | 1 | 0 | 1 | 1 | 0 | 1 | 1 | 7 |
| Arentz 2022 | 1 | 1 | 1 | 1 | 0 | 1 | 1 | 0 | 1 | 1 | 8 |
| Filardo 2022 | 1 | 1 | 1 | 1 | 0 | 1 | 1 | 1 | 1 | 1 | 9 |
| Gandhi 2022 | 0 | 1 | 0 | 1 | 1 | 1 | 1 | 1 | 1 | 1 | 8 |
| Geric 2021 | 1 | 1 | 0 | 1 | 0 | 1 | 1 | 1 | 1 | 1 | 8 |
| Godoy 2022 | 0 | 1 | 0 | 1 | 0 | 1 | 1 | 0 | 1 | 1 | 6 |
| Golandaj 2022 | 1 | 1 | 1 | 1 | 0 | 1 | 1 | 0 | 1 | 1 | 8 |
| Hasan 2022 | 1 | 1 | 1 | 1 | 0 | 1 | 1 | 0 | 1 | 1 | 8 |
| Soko 2020 | 0 | 1 | 1 | 1 | 1 | 1 | 0 | 1 | 1 | 1 | 8 |
| Crowder 2021 | 1 | 1 | 0 | 1 | 1 | 1 | 0 | 1 | 1 | 1 | 8 |
| Lungu 2022 | 1 | 1 | 0 | 0 | 1 | 1 | 1 | 0 | 1 | 1 | 7 |
| Pelissari 2022 | 1 | 0 | 0 | 0 | 1 | 1 | 1 | 0 | 1 | 1 | 6 |
| Ranasinghe 2022 | 1 | 1 | 0 | 1 | 0 | 1 | 1 | 0 | 1 | 1 | 7 |
| Summary total for each question. | 13 | 25 | 5 | 22 | 18 | 25 | 28 | 22 | 31 | 31 | |

NB

I. Was the study’s target population a close representation of the national population's relevant variables, e.g. age, sex and occupation?

I. Was the sampling frame a true or close representation of the target population?

II. Was some form of random selection used to select the sample, OR, was a census undertaken?

III. Was the likelihood of non-response, or is bias minimal?

IV. Were data collected directly from the subjects (as opposed to a proxy)?

V. Was an acceptable case definition used in the study?

VI. Was the study an instrument that measured the parameter of interest (e.g. prevalence of low back pain) shown to have reliability and validity (if necessary)?

VII. Was the same mode of data collection used for all subjects?

VIII. Was the length of the shortest prevalence period for the parameter of interest appropriate?

IX. Were the numerator(s) and denominator(s) for the parameter of interest appropriate?

X. Summary of the overall Risk of bias for each study

Yes=1

No=0

**Supplementary Table 3: Changes in TB diagnostic and treatment delay contact tracing, positivity and detection rate.**

| Study | Countries | Diagnostic delay | Treatment delay | Detection rate | Positivity rate | Contact tracing |
|---|---|---|---|---|---|---|
| Gennaro 2021 | Italy (Rome) | 45days increase between March 2019 to August 2020. | Not reported | Not reported | Not reported | Not reported |
| Gandhi 2022 | India (Northern India) | 35 days increase between January to June 2020. | 6 days increase between January to June 2020. | Not reported | Not reported | Not reported |
| Godoy 2022 | Spain (Catalonia) | Not reported | Not reported | Not reported | Not reported | 36.1% reduction during the pandemic period. |
| Mbithi 2021 | Kenya (Nairobi) | Not reported | Not reported | Not reported | 0.1% increase between March 2019 to February 2020 and March 2020 to February 2021. | Not reported |
| Srivastava | India (Gurgaon) | Not reported | Not reported | Not reported | 24.9% decrease between March 2019 to December 2019 and January 2020 to October 2020. | Not reported |

| | | | | | | |
|---|---|---|---|---|---|---|
| Thekkur 2021 | Malawi (Lilongwe) | Not reported | Not reported | Not reported | 4.5% increase between March 2019 and February 2020 to March 2020 and February 2021. | Not reported |
| Thekkur 2021 | Zimbabwe (Harare) | Not reported | Not reported | Not reported | 2.4% increase between March 2019 and February 2020 to March 2020 and February 2021. | Not reported |
| Arega 2022 | Ethiopia (Addis Ababa) | Not reported | Not reported | 11.8% and 44.7% decrease between April 2019 to March 2020 and April 2020 to March 2021. | Not reported | Not reported |

**Supplementary Table 4: Changes in TB outpatients, new cases and discharged inpatients**

| Study | Country | TB outpatients | TB new cases | TB discharged inpatients |
|---|---|---|---|---|
| Migliori 2020 | Australia | 22.1% increase between January-April 2019 and January-April 2020. | 48.6% increase between January-April 2019 and January-April 2020. | 20.7% decrease between January-April 2019 and January-April 2020. |
| Migliori 2020 | India | 71.6% decrease between January-April 2019 and January-April 2020. | Not reported | 63% decrease between January-April 2019 and January-April 2020. |
| Migliori 2020 | Philippines | 66.7% decrease between January-April 2019 and January-April 2020. | 71.4% increase between January-April 2019 and January-April 2020. | 6.1% decrease between January-April 2019 and January-April 2020. |

| | | | | |
|---|---|---|---|---|
| Migliori 2020 | Singapore | 17.5% increase between January-April 2019 and January-April 2020. | 48.4% decrease between January-April 2019 and January-April 2020. | 12.1% increase between January-April 2019 and January-April 2020. |
| Migliori 2020 | France | 40.1% increase between January-April 2019 and January-April 2020. | 31.4% decrease between January-April 2019 and January-April 2020. | 12.9% decrease between January-April 2019 and January-April 2020. |
| Migliori 2020 | Italy | 17.1% decrease between January-April 2019 and January-April 2020. | 4.8% decrease between January-April 2019 and January-April 2020. | 13.3% decrease between January-April 2019 and January-April 2020. |
| Migliori 2020 | Netherlands | Not reported | 46.0% decrease between January-April 2019 and January-April 2020. | 5.4% decrease between January-April 2019 and January-April 2020. |
| Migliori 2020 | Russia | 10.3% decrease between January-April 2019 and January-April 2020. | 11.1% decrease between January-April 2019 and January-April 2020. | 31.3% decrease between January-April 2019 and January-April 2020. |
| Migliori 2020 | Spain | 1.0% increase between January-April 2019 and January-April 2020. | 31.3% decrease between January-April 2019 and January-April 2020. | 41.7% decrease between January-April 2019 and January-April 2020. |
| Migliori 2020 | UK | 1.1% decrease in TB outpatients between January-April 2019 and January-April 2020. | 6.3% increase between January-April 2019 and January-April 2020. | 42.9% decrease between January-April 2019 and January-April 2020. |
| Migliori 2020 | Mexico | 43.2% decrease between January-April 2019 and January-April 2020. | 47.5% decrease between January-April 2019 and January-April 2020. | 90.8% decrease between January-April 2019 and January-April 2020. |
| Migliori 2020 | Argentina | 3.9% decrease between January-April 2019 and January-April 2020. | 2.6% decrease between January-April 2019 and January-April 2020. | Not reported |
| Migliori 2020 | Brazil | 0.5% decrease between January-April 2019 and January-April 2020. | 20.5% decrease between January-April 2019 and January-April 2020. | 24.0% decrease between January-April 2019 and January-April 2020. |
| Migliori 2020 | Kenya (Nairobi) | Not reported | There was a 12.6% decrease in TB new cases between January to April 2019 and 2020. | Not reported |

| | | | | |
|---|---|---|---|---|
| Migliori 2020 | Niger | 15.6% decrease between January-April 2019 and January-April 2020. | 15.6% decrease between January-April 2019 and January-April 2020. | Not reported |
| Migliori 2020 | Sierra Leone (Free Town) | 30.1% decrease between January-April 2019 and January-April 2020. | 26.5% decrease between January-April 2019 and January-April 2020. | Not reported |
| Srivastava 2021 | India (Gurgaon) | Not reported | Not reported | 15.4% increase between March 2019 to October 2020. |

**Supplementary Table 5: Changes in sensitive TB, RR/MDR TB rate and TB Deaths**

| Study | Country | Sensitive TB | RR/MDR rate | TB deaths |
|---|---|---|---|---|
| Srivastava 2021 | India (Gurgaon) | 12.3% increase between March 2019 to October 2020. | 9.9% decrease between March 2019 to December 2019 and January 2020 to October 2020. | 2.6% increase between March 2019 to October 2020. |
| Dara 2021 | 48 European countries | Not reported | 33.5% decrease between January to June 2019 and January to June 2020. | Not reported |
| Arega 2022 | Ethiopia (Addis Ababa) | Not reported | 27.7% increase between April 2019 to March 2020 and April 2020 to March 2021. | Not reported |
| Geng 2021 | China (Henan province) | Not reported | 5.1% increase in the MTB cumulative rate between January to December 2019 and January to December 2020. | Not reported |
| Mbithi 2021 | Kenya (Nairobi) | Not reported | Not reported | 0.8% increase between March 2019 to February 2020 and March 2020 to February 2021. |
| Thekkur 2021 | Malawi (Lilongwe) | Not reported | Not reported | 0.6% decrease between March 2019 and February 2020 to March 2020 and February 2021. |

| | | | | |
|---|---|---|---|---|
| Lakoh 2021 | Sierra Leone (Free Town) | Not reported | Not reported | 51.4% decrease between January 2019 to September 2019 and January 2020 to September 2020. |
| Hasan 2022 | Vietnam (all provinces) | Not reported | 1.3% decrease between 2018 and 2020. | 67% decrease between January 2019 to December 2019 and January 2020 to December 2020. |
