## Supplementary Doc for "Impact of COVID-19 on the cascade of care for tuberculosis: A systematic review"

**Supplementary document 1**

**Search strategy table**

| S/N | Search |
| --- | --- |
| #1 TB | Tuberculosis OR tuberculosis OR “Mycobacterium tuberculosis Infection” OR TB OR “active TB” OR “symptomatic TB” OR “asymptomatic TB” OR “latent TB” OR “MDR tuberculosis” OR “multidrug resistance TB” OR “XDR tuberculosis” OR “Extensive drug resistance TB” OR “MDR-TB” OR “multidrug resistance tuberculosis” OR “XDR-TB” OR “Extensive drug resistance tuberculosis” OR “tuberculosis infection” OR “TB infection” OR “Pulmonary tuberculosis” OR “Mycobacterium tuberculosis” Filters: from 2018-2021 |
| #2 COVID-19 | COVID19 OR covid19 OR coronavirus OR SARS OR sars OR severe acute respiratory syndrome OR covid19 OR cov2 OR COV2 OR “2019-nCoV” OR “SARS-CoV-2” OR “Severe Acute Respiratory Coronavirus 2” OR “coronavirus infections” OR “bat coronavirus” OR "betacoronavirus 1" OR "betacoronavirus" OR “coronavirus disease 2019” OR “Coronavirus Infection” OR Coronaviruses OR “nCoV” OR “Coronavirus Infection Disease 2019” OR “Novel Coronavirus Pneumonia” OR “2019-nCoV Infections” OR “2019 novel coronavirus” OR “2019 novel coronavirus infection” Filters: from 2018 – 2021 |
| #3 TB AND COVID-19 | (#1) AND (#2) |
